# Cost-effectiveness of a package to ensure safe prescription of 7-day high-dose primaquine for the radical cure of *Plasmodium vivax* in Indonesia

**DOI:** 10.64898/2026.09.23.26363845

**Authors:** Patrick Abraham, Liony Fransisca, Jacklyn Adella, Annisa Rahmalia, Vincent Jimanto, Sherley Angeline, Adela Putri, Kylie Mannion, Vanessa S Sakalidis, Inge Sutanto, Ayodhia Pitaloka Pasaribu, Jeanne Rini Poespoprodjo, Julie A. Simpson, Ric N. Price, Angela Devine

## Abstract

The radical cure of *Plasmodium vivax* requires treatment of the dormant liver stages with either primaquine or tafenoquine, but widespread implementation of this policy is constrained by the risk of severe hemolysis in patients with glucose-6-phosphate dehydrogenase (G6PD) deficiency. A revised case-management package, ‘New Primaquine Treatment’ (NPT), was designed to support safe radical cure in Indonesia. The package includes G6PD screening, targeting high-dose primaquine and an early clinical review to encourage full adherence to a complete course of treatment providing no early signs of adverse reactions. A decision-tree model was constructed using individual-level data from a study that treated patients with 7mg/kg primaquine according to their G6PD activity: primaquine administered over 7 days (G6PD normal individuals), 14 days (individuals with intermediate G6PD deficiency), or 8 weeks (if severely G6PD deficient). The model incorporated costs, adverse events, and the risk of *P. vivax* recurrence over a six-month time horizon. A business-as-usual comparator was created using historical *P. vivax* recurrence data for Indonesia overall and the three provinces separately. The primary outcome was disability-adjusted life-years (DALYs) averted. Costs were analyzed from health system and societal perspectives, and incremental cost-effectiveness ratios (ICERs) were compared with Indonesia’s Health Technology Assessment recommended willingness-to-pay thresholds of 1 and 3 gross domestic product per capita (US$4,925 and$14,475). One-way and probabilistic sensitivity analyses were conducted to assess uncertainty. Overall, NPT averted 0.0011 DALYs per patient with an incremental cost of US$7.40 per patient from a health system perspective. This resulted in an ICER of US$6,636 per DALY averted which was not cost-effective at the $4,925 threshold. Cost-effectiveness varied across settings: from US$5,098 per DALY averted in Papua to US$20,051 per DALY averted in southern Sumatra and US$80,825 per DALY averted in northern Sumatra, largely due to varied endemicity and case numbers. However, when the societal perspective was included, the ICER reduced significantly resulting in NPT being cost-effective overall in Indonesia ($2,261/DALY averted), and this was particularly apparent in highly endemic Papua ($675/DALY averted). The risk of *P. vivax* recurrence and intervention costs were the most influential parameters on the ICER. Overall, NPT fell between the 1-3x GDP per capita cost-effectiveness thresholds in Indonesia; however, cost-effectiveness varied significantly across settings, indicating that NPT should be prioritized for areas with high incidence of *P. vivax* malaria.

## INTRODUCTION

*Plasmodium vivax* is the predominant cause of malaria outside of Africa[1, 2], causing substantial health and economic burden in affected populations. *P. vivax* is more difficult to treat and eliminate than *P. falciparum* because it forms dormant liver stages (hypnozoites) that can reactivate months after the initial infection, causing recurrent malaria episodes (relapses), a cumulative risk of anemia, and ongoing transmission of the parasite.[1, 4, 5] Recurrent infections account for up to 85% of all blood-stage infections[6]; hence, the successful treatment of patients with *P. vivax* requires clearance of both the blood and liver stages of the parasite[7], a combination known as radical cure. Radical cure of *P. vivax* remains a significant challenge for the control and ultimate elimination of the parasite.[8] The 8-aminoquinoline compounds, primaquine and tafenoquine, are the only licensed drugs for the treatment of *P. vivax* liver stages.[9]

The efficacy of primaquine is related to the total dose administered; with high-dose regimens (7.0mg/kg total dose) reducing the risk of recurrent parasitemia by almost 50% in most endemic settings compared to low-dose regimens (3.5mg/kg total dose).[10, 11] Primaquine and tafenoquine can both induce severe hemolysis in individuals with glucose-6-phosphate-dehyrdogenase (G6PD) deficiency[12], an inherited enzymopathy present in up to 30% of malaria-endemic populations.[13, 14] Patients with ≥70% enzyme activity are considered G6PD normal, those with 30-70% activity are considered intermediate, and those with less than 30% activity are considered to be deficient. Although the World Health Organization (WHO) recommends testing patients for G6PD deficiency prior to prescribing primaquine or tafenoquine, in practice this is often not available.[15] Many malaria control programs have therefore opted for a low total dose of primaquine (3.5mg/kg) to reduce the risk of hemolysis in the absence of G6PD screening.[7]

The current Indonesian guidelines for *P. vivax* treatment recommend low-dose primaquine (0.25 mg/kg per day) administered over 14 days, totaling 3.5 mg/kg, when G6PD status is unknown.[16] Whilst this conservative approach may reduce the risk of drug-induced hemolysis, the extended length of treatment is associated with suboptimal adherence as patients often do not adhere to a complete dose of radical cure.[17, 18] Effectiveness of low-dose primaquine in routine clinical practice in Papua has been estimated to be as low as 14%.[19]

To improve the effectiveness of primaquine radical cure, a short-course high daily dose regimen has been proposed, in which patients are prescribed a high total dose regimen (7mg/kg) administered over 7 days (PQ7). PQ7 has been shown to have the same efficacy as those treated with the same total dose over 14 days.[20, 21] PQ7 requires a higher daily dose (1mg/kg/day), and this increases the risk of severe drug-induced hemolysis in patients with G6PD deficiency.[22] As a result, the WHO endorsed the use of PQ7 in 2024, but restricted it to patients with >70% G6PD activity.[14]

Malaria endemicity in Indonesia is diverse,[23] with the provinces in Java island having already achieved or near elimination,[24] whilst other provinces in Sumatra and Kalimantan have achieved very low endemicity.[25] Conversely, in the eastern Indonesian province of Papua, *P. falciparum* and *P. vivax* are both highly prevalent with recurrent episodes of *P. vivax* associated with cumulative risk of severe anemia, and its associated morbidity and mortality.[26]

Indonesia has set a national target of malaria elimination by 2030. To eliminate *P. vivax*, policymakers need to allocate resources efficiently and balance the risk of recurrence with the risk of drug-induced hemolysis across these vastly different settings. Recent advances in point-of-care diagnostics offer great potential to identify patients at greatest risk of hemolysis so that treatment can be tailored to an individual’s G6PD status, according to recent WHO recommendations.[14]. The STANDARD G6PD Test (SD Biosensor) is a semi-quantitative assay to determine an individual’s G6PD activity at the point-of-care[15], however, requires additional health systems costs for testing, equipment, and training.[27–30]

The Short COurse PrimaquinE for the radical cure of *P. vivax* (SCOPE) implementation study[31] aimed to investigate the safety, feasibility, and cost-effectiveness of an intervention package called ‘New Primaquine Treatment’ (NPT) for patients with *P. vivax* malaria in Indonesia, consisting of high-dose primaquine with quantitative G6PD testing, education, and community follow-up on day 3 from a community health worker.[32] The study took place in six community health clinics (“puskesmas”) in Indonesia, four in highly endemic Papua, one in northern Sumatra, and one in southern Sumatra. In this analysis, we explore the cost-effectiveness of the intervention package including G6PD screening before the prescription of PQ7 to those testing G6PD normal, compared to the current low-dose primaquine regimen in Indonesia.

## MATERIALS AND METHODS

### Study Design

A decision tree model was constructed in TreeAge Pro (R2.0) over a six-month time horizon using patient-level data on costs, adverse events, and G6PD status from the study data from the NPT intervention.[31] The primary analysis was performed from a health system perspective, with a scenario analysis from the societal perspective. NPT categorized patients with a *P. vivax* infection into three groups depending on their G6PD activity: patients were treated with 1mg/kg/day primaquine for 7 days if their G6PD activity was >70%, 0.5mg/kg/day for 14 days if their G6PD activity was between 30-70%, and 0.75 mg/kg primaquine per week for 8 weeks if their G6PD activity was <30%.[31, 32]

A business-as-usual comparator was created using historical risk of recurrence data from study sites[33, 34]; in this scenario, patients with *P. vivax* were assumed to have been treated with the standard low-dose primaquine regimen (3.5 mg/kg total dose) administered over 14 days, as per current standard treatment guidelines in Indonesia,[35] and this reflects current very low adherence in these settings. Pregnant or breastfeeding women, children under 6 months and patients under 5kgs were not treated with NPT in line with Indonesian national guidelines. Patients with hemoglobin <8 g/dL were also excluded from high-dose treatment. The model structure was constructed using estimates of adverse events and G6PD status parameters derived from the SCOPE study populations at each site (Figure 1), as well as historical safety data from the same locations.[33] Data for the model that were not available from the study were derived from the best available evidence from the literature (Table 1). The Consolidated Health Economic Evaluation Reporting Standards 2022 reporting guidelines for economic evaluation were adhered to (Table A in S1 Text).[36]

**Figure 1.**
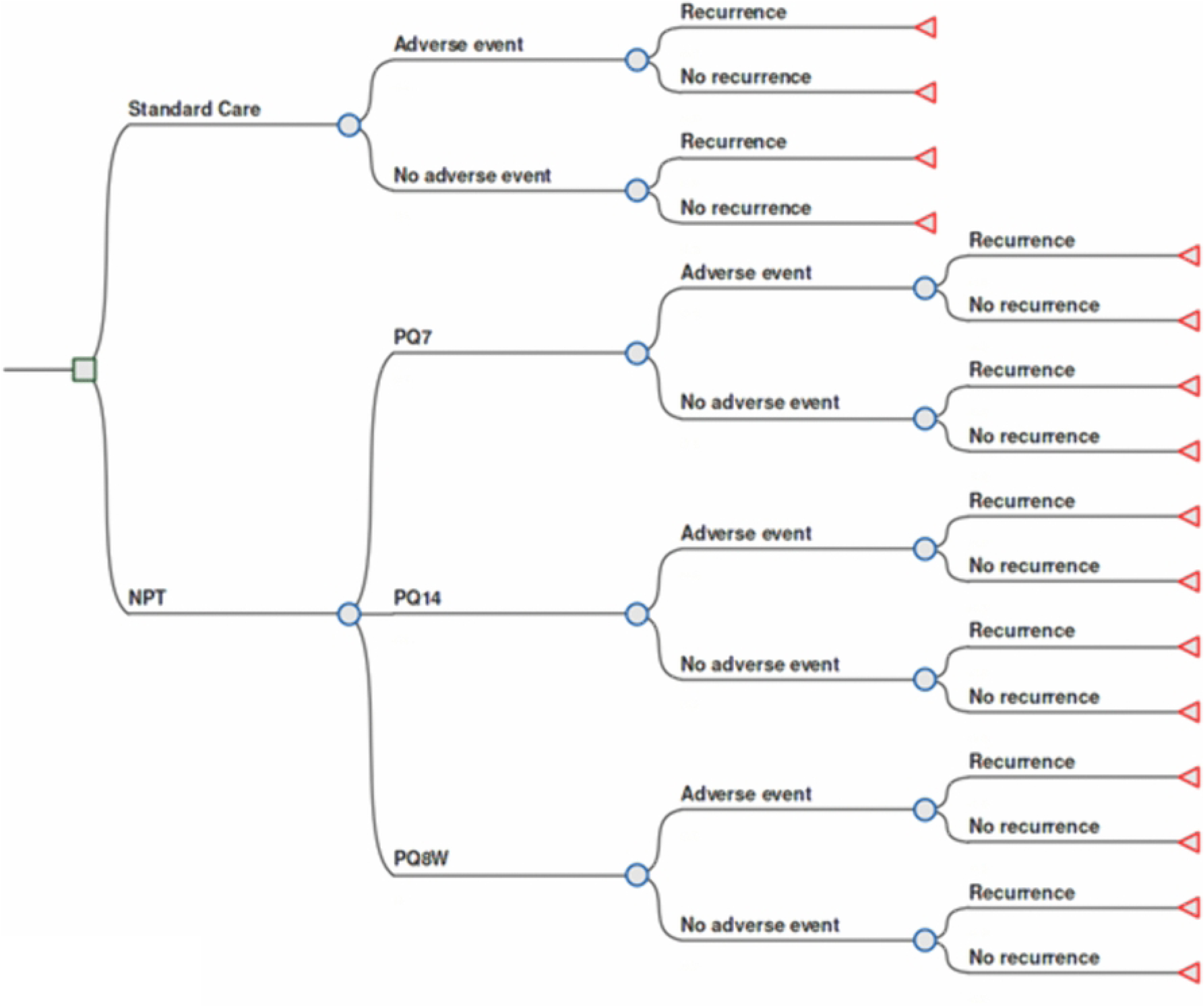
Simplified model structure for patients who are not contraindicated for primaquine treatment (e.g., pregnant or breastfeeding women, children under 6 months, patients with hemoglobin <8 g/dL). NPT: New Primaquine Treatment; PQ7: Short-course high-dose primaquine for 7 days (total dose 7mg/kg); PQ14: high-dose primaquine for 14 days (total dose 7mg/kg); PQ8W: weekly primaquine doses over 8 weeks (total dose 6mg/kg)

**Table 1.** Location of parameter values and data sources used in the cost-effectiveness model.

| Category | Where detailed |
| --- | --- |
| *Health outcomes* |  |
| Recurrences | Tables 2 & 3 |
| Adverse events | Table 3 |
| Mortality rates | Table 2 |
| Life expectancy | Table 2 |
| Disability weights | Table 2 |
| *Population parameters* |  |
| Age | Table 3 |
| G6PD status | Table 3 |
| Detailed patient demographics | Table B in S1 Text |
| *STANDARD G6PD Biosensor* |  |
| Diagnostic accuracy | Table 2 |
| *Costs* |  |
| Healthcare provider costs | Table 2 |
| Intervention costs | Table 3 & Table C in S1 Text |
| Productivity losses | Table 3 & Tables D & E in S1 Text |
| Out-of-pocket costs | Table 3 & Table E in S1 Text |

G6PD: Glucose-6-phosphate dehydrogenase

The risks of *P. vivax* recurrence 6 months after initial treatment were derived by passive detection of cases, by linking patients re-presenting to the same clinic with a further episode of *P. vivax* malaria. The risks of recurrence following standard care could not be accurately established due to significant ascertainment bias in detecting recurrence between patients enrolled into the original study (who were treated with NPT), and those not enrolled in the study (who were treated with the standard low-dose primaquine regimen). Therefore, estimates of the risk of recurrence following low-dose primaquine were derived from published trials. In Papua, the 6-month cumulative risks of patients with recurrent *P. vivax* were derived from a recent trial of patients treated with high-dose unsupervised primaquine[33] and patients enrolled and treated with low-dose primaquine (unsupervised except for a day 3 review) in Sumatra[34] (Table 2). These parameters were also considered in a scenario analysis using adjusted hazard ratios derived from a systematic review and individual patient data meta-analysis of Indonesian patients treated with high-dose primaquine compared to low-dose primaquine.[11]

**Table 2.** Cost-effectiveness model parameters taken from literature. All costs are in 2024 United States dollars.

|  | Value (Range) | Distribution | Source |
| --- | --- | --- | --- |
| *Recurrences* |  |  |  |
| Cumulative risk of recurrence at 6 months in Papua | 0.471 (0.339 – 0.625) | Beta | [33] |
| Cumulative risk of recurrence at 6 months in Sumatra | 0.050 (0.007 – 0.305) | Beta | [34] |
| Cumulative risk of recurrence at 6 months overall | 0.399 (0.282 – 0.570) | Beta | [33] [34] weighted between sites by participant number |
| Adjusted hazard ratio of recurrence, high-dose compared to low-dose (scenario analysis only) | 0.53 (0.45 – 0.63) | Beta | [11] |
| Diagnostic accuracy of the STANDARD G6PD Biosensor at 70% threshold |  |  |  |
| Sensitivity | 0.925 (0.875 – 0.975) | Beta | [37] |
| Specificity | 0.950 (0.900 – 1.00) | Beta | [37] |
| *Length of illness in days* |  |  |  |
| Anemia due to malaria | 30 (15 – 60) | Beta | [38] |
| Gastrointestinal event | 3 (1 – 7) | Beta | Assumption |
| Methemoglobinemia | 7 (3 – 10) | Beta | Assumption |
| *Disability Weights* |  |  |  |
| Malaria | 0.051 (0.032 – 0.074) | Gamma | [39] |
| Severe malaria | 0.133 (0.088 – 0.190) | Gamma | [39] |
| Anemia | 0.052 (0.034 – 0.076) | Gamma | [39] |
| Gastrointestinal event | 0.114 (0.078 – 0.159) | Gamma | [39] |
| Methemoglobinemia | 0.052 (0.034 – 0.076) | Gamma | [39] |
| *Life expectancy* |  |  |  |
| 20 – 24-year-old male | 50.3 (32.1 – 67.2) | Triangle | [40] |
| 20 – 24-year-old-male discounted* | 25.8 (20.6 – 28.9) | Triangle | [40] |
| 20 – 24-year-old female | 55.2 (36.3 – 72.6) | Triangle | [40] |
| 20 – 24-year-old female discounted* | 26.8 (22.1 – 29.6) | Triangle | [40] |
| *Costs* |  |  |  |
| Provider cost per clinic visit | \$15.90 (11.4 – 20.4) | Gamma | [41] |
| Provider cost per hospitalization for malaria | \$195.40 (162.1 – 228.7) | Gamma | [41] |
| Provider cost per hospitalization for hemolysis | \$198.06 (99.09 – 297.15) | Gamma | 7 day inpatient stay [42] |
| *Mortality* |  |  |  |
| Probability of death due to hemolysis | 0.005 (0.001 – 0.010) | Beta | [43] |
| Probability of death due to malaria | 0.00322 (0.00241 – 0.00456) | Beta | [44] |

\*Life years discounted at 3%

### Resource use and costs

Except for the costs of outpatient and inpatient visits (Table 2), all resource use and cost data were collected alongside NPT implementation. Household costs and productivity losses were collected from patients with *P. vivax* malaria attending the six study clinics pre- and post-NPT intervention, using structured, pre-tested, household economic surveys.[45] Household costs and productivity losses included data on out-of-pocket spending, lost income, and proxies of monthly spending when the income for informal workers could not be estimated. The collected data were converted to daily values and multiplied by the number of days that patients or their carers were unable to undertake usual activities (supplementary material). For costs collected in previous years or in Indonesian Rupiah, World Bank gross domestic product (GDP) deflators[46] and exchange rates[47] were applied to convert to 2024 United States dollars ($). Analysis of patient-level data for costs and length of illness was performed in Stata SE, version 18.1 (StataCorp).

Health facility costs of *P. vivax* treatment were estimated from an analysis of the costs of malaria treatment from the National Health Insurance claims dataset.[41] The health systems costs for NPT included additional primaquine, G6PD screening, staff training, and a day 3 review with a healthcare worker. These costs were collected from study records and consultation with the Indonesian Ministry of Health, with details presented in Table C in S1 Text.

### Health outcomes

The primary outcome for the cost-effectiveness analysis was disability-adjusted life-years (DALYs) due to malaria episodes (including recurrent episodes) and adverse events attributable to primaquine. The latter included severe hemolysis, gastrointestinal events and methemoglobinemia, and the risk of these adverse events associated with NPT. Numbers of these events are presented in Table 2. The length of malaria illness was taken from study surveys (Table D in S1 Text). Mortality attributable to malaria was derived from a previous study in Papua,[44] and mortality attributable to severe hemolysis was derived from a previous study in G6PD-deficient children.[43] Years of life lost were calculated using Indonesian life tables[48] and the median age of study participants (Table B in S1 Text).

### Analysis

The ICERs were calculated by dividing the incremental cost of the treatment package by the DALYs averted as follows:

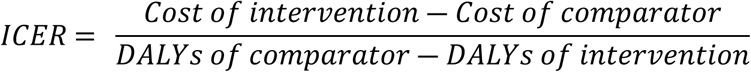

As per the Indonesian Health Technology Assessment Guidelines,[49] two willingness to pay (WTP) thresholds were used: US$4,925 and US$14,775, representing 1x and 3x Indonesia’s GDP per capita.[50] One-way sensitivity analyses of epidemiologic, cost, and safety parameters were performed using parameter ranges from Tables 2 and 3. A two-way sensitivity analysis considered a range of values for both the additional cost and the risk of recurrence for NPT to evaluate whether standard care or NPT were cost-effective at the 1x GDP per capita threshold.

**Table 3.** Overall and site-specific model parameters and ranges taken directly from NPT data along with the distributions used in the probabilistic sensitivity analysis. All costs are in 2024 United States dollars.

|  | Overall | Papua | Southern Sumatra | Northern Sumatra | Distribution |
| --- | --- | --- | --- | --- | --- |
| Proportion of patients who are G6PD intermediate | 0.124 (0.116 – 0.136) | 0.117 (0.105 – 0.129) | 0.234 (0.211 – 0.257) | 0.111 (0.100 – 0.122) | Beta |
| Proportion of patients who are G6PD deficient | 0.019 (0.017 – 0.021) | 0.020 (0.018 – 0.022) | 0.016 (0.014 – 0.018) | 0.008 (0.007 – 0.009) | Beta |
| Cumulative risk of recurrence at 6 months with NPT | 0.166 (0.148 – 0.185) | 0.203 (0.181 – 0.226) | 0.034 (0.019 – 0.059)* | 0.034 (0.019 – 0.059)* | Beta |
| *Costs* |  |  |  |  |  |
| NPT | \$13.34 (6.67 – 20.01) | \$12.69 (6.34 – 19.03) | \$12.96 (6.48 – 19.44) | \$39.90 (19.95 – 59.85) | Gamma |
| Out-of-pocket costs | \$4.75 (0.00 – 41.35) | \$2.80 (0.05 – 10.96) | \$7.87 (0.00 – 51.20) | \$7.90 (0.01 – 37.83) | Gamma |
| Productivity losses | \$16.11 (0.00 – 125.28)) | \$11.98 (0.00 – 121.14) | \$18.46 (0.03 – 88.20) | \$26.73 (0.01 – 138.47) | Gamma |
| *Length of illness (days)* |  |  |  |  |  |
| Malaria | 4.9 (0.8 – 9.0) | 4.9 (0.8 – 9.0) | 4.9 (0.8 – 9.0) | 4.9 (0.8 – 9.0) | Gamma |
| *Risk of adverse events by treatment regimen* |  |  |  |  |  |
|  | All | PQ7 | PQ14 | PQ8W |  |
| Hemolysis | 0.005 (0.003 – 0.008) | 0.005 (0.002 – 0.007) | 0.003 (0.003 – 0.006) | 0.031 (0.031 – 0.062) | Triangle |
| Gastrointestinal | 0.006 (0.001 – 0.007) | 0.007 (0.001 – 0.008) | 0.003 (0.000 – 0.003) | 0 (0 – 0) | Triangle |
| Methemoglobin | 0.001 (0.0004 – 0.002) | 0.001 (0.0004 – 0.002) | 0 (0 – 0) | 0 (0 – 0) | Triangle |

G6PD: Glucose-6-phosphate dehydrogenase; NPT: New Primaquine Treatment, PQ7: Short-course high-dose primaquine for 7 days (total dose 7mg/kg); PQ14: high-dose primaquine for 14 days (total dose 7mg/kg); PQ8W: weekly primaquine doses over 8 weeks (total dose 6mg/kg)
\*Risk of recurrence estimates were pooled in Southern and Northern Sumatra due to small patient numbers

**Table 4.** Cost-effectiveness of NPT compared to standard care in Indonesia overall, Papua, southern Sumatra, and northern Sumatra, from the health system and societal perspectives in 2024 United States dollars.

| Location | DALYs | DALYs averted | Health system perspective |  |  | Societal perspective |  |  |
| --- | --- | --- | --- | --- | --- | --- | --- | --- |
|  |  |  | Cost | Incremental cost | ICER | Cost | Incremental cost | ICER |
| *Indonesia overall* |  |  |  |  |  |  |  |  |
| Standard Care | 0.0065 | base | \$24.50 | base | base | \$53.74 | base | base |
| NPT | 0.0054 | 0.0011 | \$31.90 | \$7.40 | **\$6,636** | \$56.26 | \$2.52 | **\$2,261** |
| *Papua* |  |  |  |  |  |  |  |  |
| Standard Care | 0.0068 | base | \$25.88 | base | base | \$56.78 | base | base |
| NPT | 0.0056 | 0.0012 | \$32.12 | \$6.23 | **\$5,098** | \$57.60 | \$0.83 | **\$675** |
| *Southern Sumatra* |  |  |  |  |  |  |  |  |
| Standard Care | 0.0052 | base | \$19.72 | base | base | \$43.14 | base | base |
| NPT | 0.0047 | 0.0005 | \$29.42 | \$9.70 | **\$20,051** | \$51.03 | \$7.89 | **\$16,298** |
| *Northern Sumatra* |  |  |  |  |  |  |  |  |
| Standard Care | 0.0051 | base | \$18.77 | base | base | \$42.19 | base | base |
| NPT | 0.0046 | 0.0004 | \$56.35 | \$37.58 | **\$80,823** | \$77.96 | \$35.77 | **\$76,917** |

DALY: Disability-adjusted life-year; ICER: Incremental cost-effectiveness ratio; NPT: New Primaquine Treatment.

Probabilistic sensitivity analyses estimated the uncertainty on the ICER estimates, using 10,000 Monte Carlo simulations. To assess uncertainty in the decision to adopt NPT, a cost-effectiveness acceptability curve was constructed by calculating the proportion of simulations which fall below a range of cost-effectiveness thresholds from $0/DALY averted to US$20,000/DALY averted.[51]

Several scenario analyses were also conducted to test the plausibility of the intervention’s cost-effectiveness to determine conditions which could guide policymakers to feasibly implement the intervention in a variety of settings. First, the analysis was conducted from a societal perspective to explore the effect of patients’ out-of-pocket spending and household lost productivity on overall results. Secondly, an additional scenario was created in which there was no mortality attributable to either hemolysis or malaria, to reflect the lack of mortality observed during the study. Lastly, to overcome uncertainty in the risk of recurrence due to ascertainment bias of passive case detection during the study, an alternate recurrence risk for NPT was generated using parameters derived from a systematic review and individual patient data meta-analysis of high-dose compared to low-dose primaquine in Indonesia (Table 2).[11]

### Ethics

The study protocol was approved by the World Health Organization Ethics Committee (Indonesia: ERC 0003810, PNG ERC 0003892), Menzies School of Health Research (HREC: 2023–4524), Alfred Health (Burnet Institute, Project No: 18/23), University of Gadjah Mada (KE/FK/0079/EC/2023), University of Indonesia (KET 347/UN2.F1/ETIK/PPM.00.02/2023), University of North Sumatra (80/KEPK/USU/2023). All participants provided written, informed consent to join the study.[32]

## RESULTS

The parameters used within the cost-effectiveness model that were generated directly from the NPT data are presented in Table 3. These include the proportion of participants within each of the model branches (based on G6PD activity), adverse events, intervention costs, productivity losses associated with malarial illness, and 6-month cumulative risk of recurrence. The cost of NPT varied widely between study sites, ranging from $12.69 per patient in Papua to $39.90 in North Sumatra. The costs were largely dependent on the case numbers per clinic and local cost factors such as costs of materials and required training (Table C in S1 Text). Reported out-of-pocket costs per patient were $4.75 (standard deviation (SD) = 8.56) and productivity losses were estimated at $16.1 (SD = 37.33) overall; site-specific estimates are presented in Table E in S1 Text.

### Base case analysis

Overall in Indonesia, NPT averted 0.0011 DALYs due to reduced recurrences at an incremental cost of $7.40 per *P. vivax* patient. From a health system perspective, this resulted in an ICER of $6,636 per DALY averted, indicating the intervention was not cost-effective at the lower WTP threshold ($4,925), but cost-effective at the higher threshold ($14,775). The cost-effectiveness results varied significantly across the three settings. In the highly endemic setting of Papua, NPT showed an ICER of $5,098 per DALY averted. In southern Sumatra, NPT was less cost-effective with an ICER of $20,051 per DALY averted, above the higher WTP threshold. NPT was also not cost-effective in northern Sumatra, largely due to high intervention costs and lower case numbers (ICER = $80,823).

From a societal perspective, the NPT was cost-effective at the lower WTP threshold in Indonesia overall and in Papua, with ICERs of $2,261 and $675 per DALY averted, respectively. While the ICERs in northern and southern Sumatra both decreased with the societal perspective, both remained above the higher WTP threshold. When considering the scenario in which there was no attributable mortality, the ICER for Indonesia overall increased to $17,628 per DALY averted, above both thresholds (Table F in S1 Text). When using an adjusted hazard ratio scenario derived from supervised treatments, the ICER for Indonesia stayed relatively stable at $6,439 (Table G in S1 Text).

### One-way sensitivity analysis

The following parameters had the greatest impact on the cost-effectiveness: i) the risk of recurrence in patients receiving NPT, ii) the discount rate applied to DALYs, iii) the cost of the intervention package, and iv) the risk of recurrence during standard care (Figure 2). High values for the discount rate and intervention cost, and the low value for the risk of recurrence under NPT caused the ICER to cross the 3x GDP threshold in all settings. The reduction in the risk of recurrence was also assessed in a one-way sensitivity analysis and demonstrated that NPT would have to reduce recurrence risk by 63% and 30% to be cost-effective at 1x and 3x GDP per capita thresholds respectively (Figure A in S1 Text).

**Figure 2.**
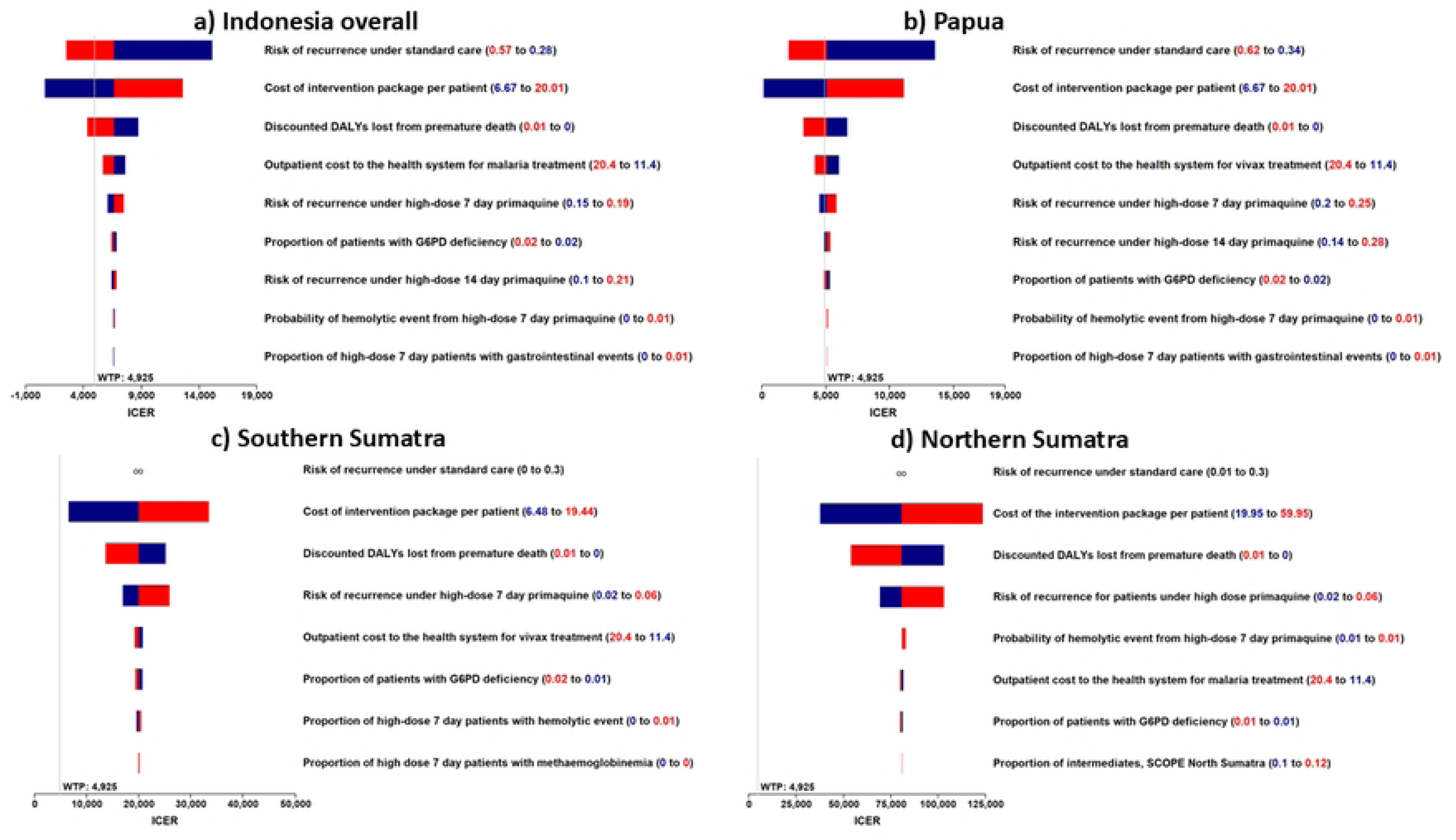
Tornado diagram of one-way sensitivity analyses indicating the nine parameters which the incremental cost-effectiveness ratio (ICER) is most sensitive in a) Indonesia overall, b) Papua c) southern Sumatra, d) northern Sumatra. The low parameter values are in blue text, while the high parameter values are in red. While the scales for Indonesia overall and Papua are the same, the scale for each Sumatran site is larger to fit uncertainty of parameters. DALYs: disability-adjusted life years; ICER: Incremental cost effectiveness ratio; G6PD: glucose-6-phosphate dehydrogenase; WTP – Willingness to Pay threshold.

### Probabilistic Sensitivity Analysis

Results from the probabilistic sensitivity analyses for Indonesia overall indicated that from a health system perspective, 33% of iterations were cost-effective at the 1x GDP per capita threshold and 88% of iterations were cost-effective at 3x GDP per capita threshold (Figure 3). The southern Sumatran site was not cost effective at either threshold; however, 33% of the model iterations were below the 3x GDP per capita threshold. In northern Sumatra, 0% of the model iterations were below the 3x GDP per capita threshold.

**Figure 3.**
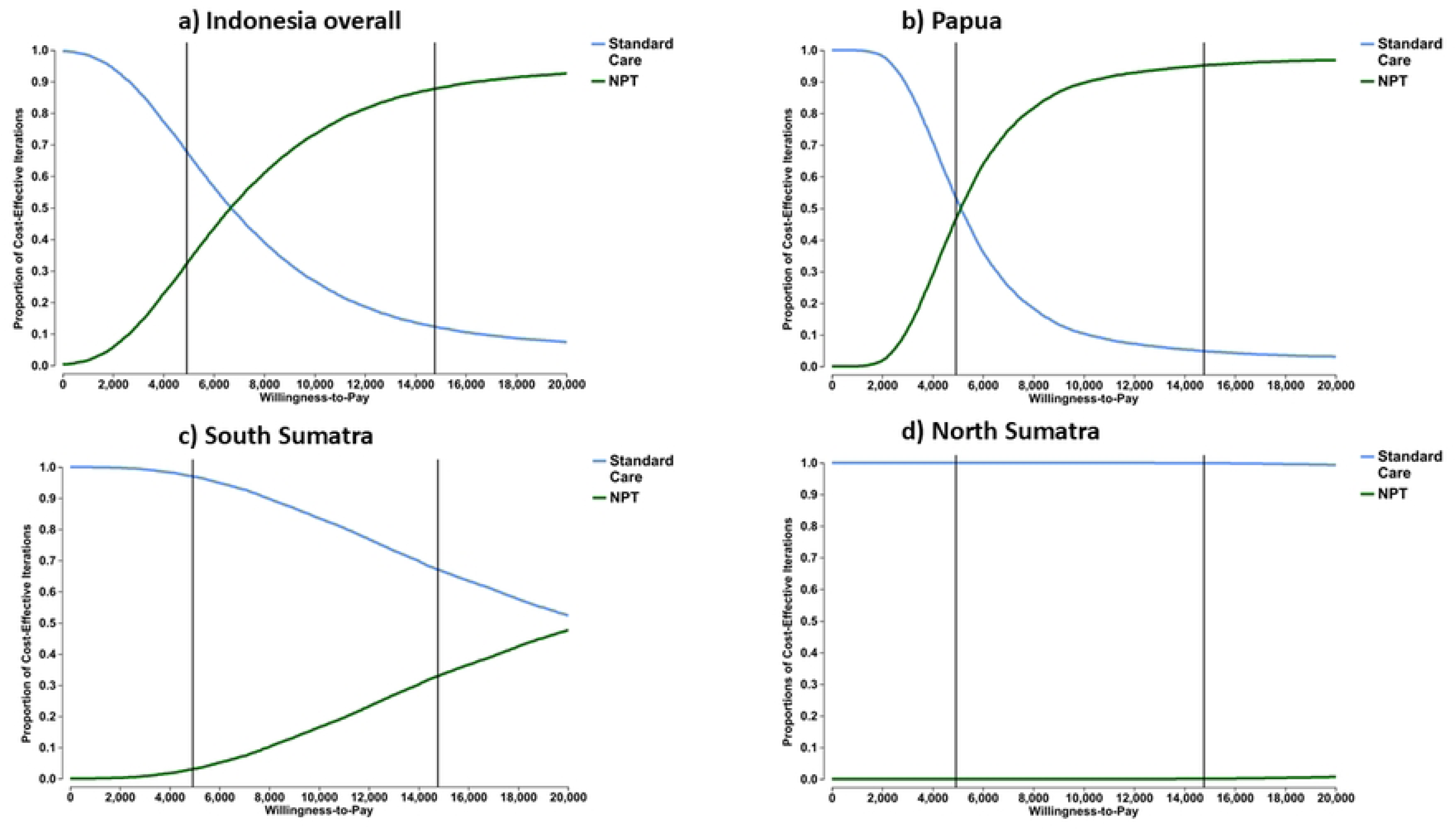
Cost-effectiveness acceptability curves from outputs of probabilistic sensitivity analyses for a) Indonesia overall, b) Papua c) southern Sumatra, d) northern Sumatra. Vertical lines indicate willingness to pay thresholds in Indonesia, representing 1 and 3x GDP per capita per DALY averted, $4,975 and $14,775, respectively, in 2024 United Stated dollars. GDP: gross domestic product; DALY: Disability adjusted life year; NPT: New Primaquine Treatment.

### Two-way sensitivity analyses

Figure 4 shows the results of the two-way sensitivity analysis, which presents a range of values for the risk of recurrence following NPT (x-axis) and the cost of the intervention per patient (y-axis). Base case parameter inputs (where black lines intersect) were close to the boundaries where business-as-usual remained cost-effective (blue region), indicating that combined uncertainty in these parameters affects the confidence in the base case ICER. At current price estimates, NPT would need to reduce the risk of recurrence by at least 50% compared to standard care, to be cost effective.

**Figure 4:**
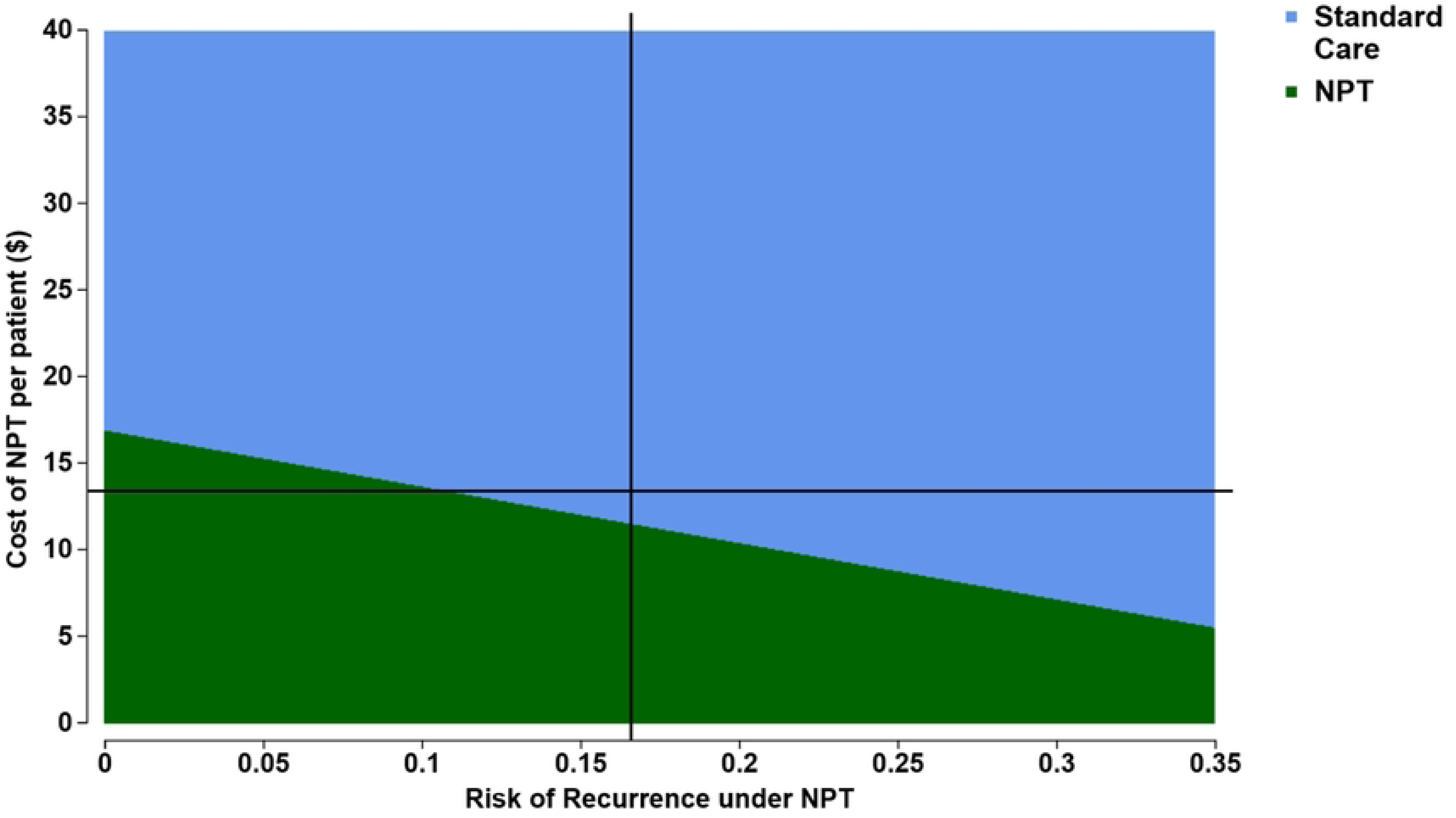
Two-way sensitivity analysis for Indonesia overall with the cost of New Primaquine Treatment (NPT) package and the 6-month risk of recurrence in patients prescribed 7-day high-dose primaquine under this package. The x-axis presents a range of plausible values for the risk of recurrent malaria episodes in patients prescribed 7-day high-dose primaquine, while the y-axis shows a range of potential costs per patient with NPT. The vertical and horizontal black lines show the base case input for these parameters. While NPT is cost effective in the parameter ranges shown in purple, standard care cost effective for parameter ranges in blue. Costs are in 2024 United States dollars. NPT: New Primaquine Treatment

## DISCUSSION

Our economic analysis is based on the first implementation study of new primaquine treatment regimen, including high-dose short-course primaquine and point-of-care G6PD testing for *P. vivax* patients[31], and provides cost-effectiveness evidence for guiding policy in Indonesia. The results demonstrate that the revised NPT case management was potentially cost effective, with the ICER falling between the 1x and 3x GDP per capita thresholds from a health system perspective. Provincial level cost-effectiveness results varied significantly between sites. In Papua the ICER was cost-effective under the lower threshold; however, in southern and northern Sumatra, NPT was not cost-effective, even when compared to the higher threshold. However, when considered from the societal perspective, the ICERs in Indonesia overall and Papua were below the lower threshold, whereas Sumatran sites remained not cost-effective. This was largely due to the significantly lower number of malaria cases and low risk of recurrence in Sumatra.

Most health economic analyses of novel radical cure strategies compare low-dose primaquine regimens with tafenoquine[29], whereas the current analysis compares the cost effectiveness of the recently WHO approved high-dose 7 day short-course (7mg/kg total dose) primaquine after G6PD screening with the low-dose 14-day primaquine regimen (3.5 mg/kg total dose), which is the current standard antimalarial treatment for patients with *P. vivax* in Indonesia. The SCOPE study demonstrated the feasibility of point-of-care G6PD testing[52] and provides important evidence to inform future considerations for adopting PQ7, and potentially tafenoquine in the future, since point-of-care testing would be required for both drug regimens.

Whilst the SCOPE study delivered a consistent implementation of NPT across different endemic settings, the cost-effectiveness analyses were substantially different. Two primary factors account for this difference: the markedly different risk of recurrence in Papua and Sumatra, and local cost factors that became influential as patient case workloads fell in pre-elimination Sumatra. Patients in Papua who are not treated with primaquine have a very high risk of *P. vivax* recurrence (>80% within 6 months) and under routine clinical practice there is only minimal reduction in relapses when patients are treated with low-dose 14-day primaquine regimen.[19, 33] Conversely, the risk of recurrence in patients infected with *P. vivax* in Sumatra is significantly lower, rarely exceeding 30% at a longer follow-up time of 12 months[21, 34] resulting a smaller reduction in DALYs as compared to Papua. Secondly, the additional costs of the G6PD test and training for health workers were averaged over the number of patients with *P. vivax* treated at each clinic. In clinics with low numbers of patients with *P. vivax,* the intervention costs per patient were significantly higher (Table C in S1 Text). The combination of these inputs resulted in the intervention not being cost-effective in either of the Sumatra sites.

Economic evaluations are designed to identify interventions that maximize population health gains by yielding the greatest health output through efficient resource allocation (allocative efficiency); however, the use of economic evaluations in malaria policy-making is debated and varied.[53] In the short term, novel malaria control interventions in settings approaching elimination are rarely cost-effective due to the high delivery costs in often hard to reach populations, meaning minimal incremental health gains from low case numbers.[53, 54] However, the long-term economic and societal benefits of even high-cost interventions can be enormous in these settings if the intervention can achieve elimination of the parasite.[55] Hence, the utility of traditional cost-effectiveness results in settings that are nearing elimination may be limited.[53]

A societal perspective is important for patients since reducing the economic burden on households has flow-on effects to economic output and financial stability for households in these settings.[56, 57] This is particularly important as the economic advantages of malarial interventions are often most beneficial to patients and families in rural and remote areas with lower financial stability.[58] Including productivity losses and out-of-pocket spending allows a broader societal perspective to be considered, and in this context the NPT intervention was more favorable across all settings. In Papua and in Indonesia overall, NPT became cost-effective at the lower threshold, indicating an economic rationale for implementation, particularly in Papua where the intervention package became highly cost-effective. However, in both Sumatran sites, the intervention was still not cost-effective at the higher 3x GDP threshold, potentially indicating the need for a more nuanced national policy that differs by province.

Our study has several limitations. Firstly, a reliable estimate of pre-and post-implementation effectiveness of NPT could not be derived directly, due to study design factors and significant ascertainment bias in consistent estimates on intervention impact. Instead, our analysis used estimates for the background recurrence risk from separate clinical trials in Papua and Sumatra.[33, 34] Since these clinical trials followed patients actively, they are less vulnerable to attrition bias when detecting recurrences compared to the passive follow-up adopted in the SCOPE study.[31] Hence, the base case analysis may have overestimated the comparative effectiveness of the NPT intervention. However, the background recurrence in Papua was based on a trial which used high-dose primaquine[33] and was therefore a conservative estimate of the true unsupervised low-dose recurrence risk. Additionally, a scenario analysis which used an adjusted hazard ratio of recurrence from a meta-analysis of high-dose and low-dose primaquine regimens[11] showed similar results for all settings, and did not change the interpretation of ICERs. The effectiveness within this scenario analysis was also assessed in a one-way sensitivity analysis, demonstrating that compared to standard care, the NPT would have to reduce the risk of recurrence by 63% to be cost-effective at 1x GDP per capita thresholds and 30% at the 3x GDP threshold (Figure A in S1 Text). This highlights the need for policy to be informed by robust estimates of the absolute difference in risk of *P. vivax* recurrence.

Secondly, we only considered relapses over a 6-month time horizon, thus longer-term impacts were not included; this approach was chosen to reflect the available effectiveness estimates of NPT. This is likely to have underestimated effectiveness since late relapses would not have been detected and also the additional impact of widespread coverage of a highly-effective primaquine regimen in reducing *P. vivax* transmission and thus incidence of malaria in the population.[59]

Lastly, the generalizability of the SCOPE study findings when the NPT intervention is scaled up beyond the selected study sites is not known. The six SCOPE sites have a rich history of undertaking clinical trials and public health research and have had substantial training. To achieve a similar level of training at less experienced health clinics would likely require significantly greater training costs. Additional study-specific effects may be reflected in the high level of safety seen for NPT; this might also vary following scale up of the intervention. Factors such as education and community engagement, which were required for NPT, are also poorly understood beyond the study sites. Future research is needed to understand how various scale-up conditions, including patient volume, training needs and health facility capacity will influence the economic feasibility of the intervention.

Future research should consider the cost of the various components of the intervention, and how representative these are of national procurement prices. The cost of the STANDARD G6PD analyzer and testing strips were derived from study records and Indonesian health resource database,[60] but these may change in the future, once taxation and procurement arrangements have been finalized. The recent volume price guarantee (US$300 per analyzer) may result in lower costs.[61] Consideration of a variety of procurement strategies will be needed to inform governments of the impact of pricing on NPT implementation.

In conclusion, the cost effectiveness of the NPT intervention varied considerably across settings, being cost-effective in highly endemic Papua from a societal perspective but not cost-effective in pre-elimination northern and southern Sumatra. Local evidence of costs and recurrence rates as well as endemicity are needed to explore the cost-effectiveness of scaling up of this intervention in other locations in Indonesia. Future research should consider local endemicity and the impact of cost inputs in a wider range of healthcare settings and locations to help determine the feasibility of Indonesia implementing NPT.

## Data Availability

The minimal dataset is available online at https://github.com/PatrickAbraham1/SCOPE_CEA.git

## Acknowledgements

This work was conducted on behalf of the Short COurse PrimaquinE for the radical cure of *P. vivax* (SCOPE Study) and funded by UNITAID. We acknowledge Medicines for Malaria Venture (MMV) as a partner and thank all collaborating institutions and field teams across Indonesia for their essential support and contributions. AD, JAS and RNP are supported by Australian National Health and Medical Research Council of Australia (NHMRC) Investigator Grants (2025362, 2042554, 2008501, respectively). This project was supported by the Australian Centre of Research Excellence in Malaria Elimination (NHMRC 2024622).

## Trial registration

The study was registered on clinicaltrials.gov, NCT05879224.

## S1 Text. Supporting information

PA was responsible for conceptualization, formal analysis, methodology, writing – original draft preparation. LF and JA were responsible for data curation, resources, writing - review and editing. AR was responsible for investigation, writing - review and editing. VJ, SA and AP were responsible for data curation, investigation and writing - review and editing. KM and VS were responsible for project administration, resources and writing - review and editing. IS, APP and JRP were responsible for conceptualization, resources, supervision and writing - review and editing. JAS, RNP were responsible for supervision, and writing - review and editing. AD was responsible for conceptualization, resources, methodology, supervision and writing - review and editing.

## REFERENCES

1. Price RN, Tjitra E, Guerra CA, Yeung S, White NJ, Anstey NM. Vivax malaria: neglected and not benign. The American journal of tropical medicine and hygiene. 2007;77(6 Suppl):79.

2. Price RN, Commons RJ, Battle KE, Thriemer K, Mendis K. Plasmodium vivax in the Era of the Shrinking P. falciparum Map. Trends in parasitology. 2020;36(6):560–70.

3. Devine A, Pasaribu AP, Teferi T, Pham HT, Awab GR, Contantia F, et al. Provider and household costs of *Plasmodium vivax* malaria episodes: a multicountry comparative analysis of primary trial data. Bulletin of the World Health Organization. 2019;97(12):828– 36. Epub 2019/12/11. doi: 10.2471/blt.18.226688. PubMed PMID: 31819291.

4. Dini S, Douglas NM, Poespoprodjo JR, Kenangalem E, Sugiarto P, Plumb ID, et al. The risk of morbidity and mortality following recurrent malaria in Papua, Indonesia: a retrospective cohort study. BMC medicine. 2020;18(1):28.

5. White NJ. Determinants of relapse periodicity in Plasmodium vivax malaria. Malaria Journal. 2011;10(1):297.

6. Commons RJ, Simpson JA, Watson J, White NJ, Price RN. Estimating the proportion of Plasmodium vivax recurrences caused by relapse: a systematic review and meta-analysis. The American journal of tropical medicine and hygiene. 2020;103(3):1094.

7. Thriemer K, Ley B, von Seidlein L. Towards the elimination of Plasmodium vivax malaria: Implementing the radical cure. PLoS medicine. 2021;18(4):e1003494.

8. Thriemer K, Bobogare A, Ley B, Gudo CS, Alam MS, Anstey NM, et al. Quantifying primaquine effectiveness and improving adherence: a round table discussion of the APMEN Vivax Working Group. Malaria Journal. 2018;17(1):241.

9. Adhikari B, Awab GR, von Seidlein L. Rolling out the radical cure for vivax malaria in Asia: a qualitative study among policy makers and stakeholders. Malaria Journal. 2021;20:1–15.

10. Commons RJ, Rajasekhar M, Edler P, Abreha T, Awab GR, Baird JK, et al. Effect of primaquine dose on the risk of recurrence in patients with uncomplicated Plasmodium vivax: a systematic review and individual patient data meta-analysis. The Lancet Infectious Diseases. 2024;24(2):172–83.

11. Fadilah I, Watson JA, Pasaribu AP, Sutanto I, Nelwan EJ, Lidia K, et al. Effect of higher dose primaquine for the radical cure of *Plasmodium vivax* malaria in Indonesia: a systematic review and individual patient data meta-analysis. The Lancet Regional Health– Western Pacific. 2026;72.

12. Ashley EA, Recht J, White NJ. Primaquine: the risks and the benefits. Malaria Journal. 2014;13(1):418.

13. Howes RE, Piel FB, Patil AP, Nyangiri OA, Gething PW, Dewi M, et al. G6PD deficiency prevalence and estimates of affected populations in malaria endemic countries: a geostatistical model-based map. PLoS medicine. 2012;9(11):e1001339.

14. World Health Organization. WHO guidelines for malaria. Geneva: World Health Organization, 2025.

15. Anderle A, Bancone G, Domingo GJ, Gerth-Guyette E, Pal S, Satyagraha AW. Point-of-care testing for G6PD deficiency: opportunities for screening. International journal of neonatal screening. 2018;4(4):34.

16. Ministry of Health of the Republic of Indonesia. Pocketbook of malaria case management. 2023.

17. Rahmalia A, Poespoprodjo JR, Landuwulang CU, Ronse M, Kenangalem E, Burdam FH, et al. Adherence to 14-day radical cure for Plasmodium vivax malaria in Papua, Indonesia: a mixed-methods study. Malaria Journal. 2023;22(1):162.

18. Mehdipour P, Rajasekhar M, Dini S, Zaloumis S, Abreha T, Adam I, et al. Effect of adherence to primaquine on the risk of Plasmodium vivax recurrence: a WorldWide Antimalarial Resistance Network systematic review and individual patient data meta-analysis. Malaria journal. 2023;22(1):306.

19. Douglas NM, Poespoprodjo JR, Patriani D, Malloy MJ, Kenangalem E, Sugiarto P, et al. Unsupervised primaquine for the treatment of *Plasmodium vivax* malaria relapses in southern Papua: A hospital-based cohort study. PLoS medicine. 2017;14(8):e1002379.

20. Chu CS, Phyo AP, Turner C, Win HH, Poe NP, Yotyingaphiram W, et al. Chloroquine versus dihydroartemisinin-piperaquine with standard high-dose primaquine given either for 7 days or 14 days in *Plasmodium vivax* malaria. Clinical Infectious Diseases. 2019;68(8):1311–9.

21. Taylor WR, Thriemer K, von Seidlein L, Yuentrakul P, Assawariyathipat T, Assefa A, et al. Short-course primaquine for the radical cure of Plasmodium vivax malaria: a multicentre, randomised, placebo-controlled non-inferiority trial. The Lancet. 2019;394(10202):929–38.

22. Thriemer K, Ley B, Bobogare A, Dysoley L, Alam MS, Pasaribu AP, et al. Challenges for achieving safe and effective radical cure of Plasmodium vivax: a round table discussion of the APMEN Vivax Working Group. Malaria Journal. 2017;16(1):1–9.

23. Herdiana H, Prameswari HD, Puspadewi RT, Fajariyani SB, Diptyanusa A, Theodora M, et al. Shrinking the malaria map in Indonesia: progress of subnational control, elimination, and future strategies. BMC medicine. 2025;23(1):512.

24. Sitohang V, Sariwati E, Fajariyani SB, Hwang D, Kurnia B, Hapsari RK, et al. Malaria elimination in Indonesia: halfway there. The Lancet Global Health. 2018;6(6):e604– e6.

25. Sugiarto SR, Baird JK, Singh B, Elyazar I, Davis TM. The history and current epidemiology of malaria in Kalimantan, Indonesia. Malaria Journal. 2022;21(1):327.

26. Fadilah I, Djaafara BA, Lestari KD, Fajariyani SB, Sunandar E, Makamur BG, et al. Quantifying spatial heterogeneity of malaria in the endemic Papua region of Indonesia: Analysis of epidemiological surveillance data. The Lancet Regional Health-Southeast Asia. 2022;5.

27. Peixoto HM, Bastos LL, Brito-Sousa JD, Sampaio VS, Daumerie PG, Jambert E, et al. The Cost-Effectiveness of Tafenoquine Following Screening with STANDARDTM G6PD Screening for the Treatment of Vivax Malaria in the Brazilian Public Health System. Available at SSRN 5151134.

28. Aung YN, Tun STT, Vanisaveth V, Chindavongsa K, Kanya L. Cost-effectiveness analysis of G6PD diagnostic test for Plasmodium vivax radical cure in Lao PDR: An economic modelling study. Plos one. 2022;17(4):e0267193.

29. Devine A. A review of the cost-effectiveness of using near-patient G6PD tests before treatment with radical cure of vivax malaria. 2025.

30. Sadhewa A, Cassidy-Seyoum S, Acharya S, Devine A, Price RN, Mwaura M, et al. A review of the current status of G6PD deficiency testing to guide radical cure treatment for vivax malaria. Pathogens. 2023;12(5):650.

31. Price RN, Robinson LJ. High-Dose, Short-Course Primaquine after Point-Of-Care G6PD Testing for the Radical Cure of Plasmodium Vivax Malaria: An Implementation Study in Papua New Guinea and Indonesia. SSRN preprint. 2026.

32. SCOPE Study Group. High daily dose Short COurse PrimaquinE after G6PD testing for the radical cure of Plasmodium vivax malaria in Indonesia and Papua New Guinea: the SCOPE implementation study protocol. BMC Infectious Diseases. 2025;25:922.

33. Poespoprodjo JR, Burdam FH, Candrawati F, Ley B, Meagher N, Kenangalem E, et al. Supervised versus unsupervised primaquine radical cure for the treatment of falciparum and vivax malaria in Papua, Indonesia: a cluster-randomised, controlled, open-label superiority trial. The Lancet Infectious Diseases. 2022;22(3):367–76.

34. Degaga TS, Pasaribu AP, Tripura R, Ghanchi N, Rajasekhar M, Adhikari B, et al. Effectiveness and safety of 7-day high-dose primaquine and single-dose tafenoquine versus 14-day low-dose primaquine in patients with *Plasmodium vivax* malaria (EFFORT): a multicentre, open-label, randomised, controlled, superiority trial. Lancet Infect Dis. 2026;26(6):614–26. Epub 20260211. doi: 10.1016/s1473-3099(25)00729-7. PubMed PMID: 41690325.

35. Kementerian Kesehatan Republik Indonesia. Pedoman Nasional Pelayanan Kedokteran (PNPK): Tata Laksana Malaria. Jakarta: Kementerian Kesehatan Republik Indonesia, 2019.

36. Husereau D, Drummond M, Augustovski F, de Bekker-Grob E, Briggs AH, Carswell C, et al. Consolidated Health Economic Evaluation Reporting Standards 2022 (CHEERS 2022) statement: updated reporting guidance for health economic evaluations. MDM Policy & Practice. 2022;7(1):23814683211061097.

37. Zobrist S, Brito M, Garbin E, Monteiro WM, Clementino Freitas S, Macedo M, et al. Evaluation of a point-of-care diagnostic to identify glucose-6-phosphate dehydrogenase deficiency in Brazil. PLoS neglected tropical diseases. 2021;15(8):e0009649.

38. Devine A, Parmiter M, Chu CS, Bancone G, Nosten F, Price RN, et al. Using G6PD tests to enable the safe treatment of Plasmodium vivax infections with primaquine on the Thailand-Myanmar border: A cost-effectiveness analysis. PLoS neglected tropical diseases. 2017;11(5):e0005602.

39. Global Burden of Disease Collaborative Network. Global Burden of Disease Study 2021 (GBD 2021) Disability Weights. Seattle, WA: Institute for Health Metrics and Evaluation (IHME); 2021.

40. Database HLT. Human Life–Table Database: Indonesia country life tables 2025. Available from: https://www.lifetable.de/Country/Country?cntr=IDN.

41. Setiawan E, Devine A, Prameswary HD, Baird JK, Price R, Thriemer K. Malaria morbidity, mortality and associated costs in Indonesia: analysis of the National Health Insurance claim dataset. BMJ Global Health. 2025;10(5).

42. World Health Organization. WHO-CHOICE estimates of cost for inpatient and outpatient health service delivery. In: Department of Health Systems Governance and Financing, editor. 2021.

43. Pamba A, Richardson ND, Carter N, Duparc S, Premji Z, Tiono AB, et al. Clinical spectrum and severity of hemolytic anemia in glucose 6-phosphate dehydrogenase–deficient children receiving dapsone. Blood, The Journal of the American Society of Hematology. 2012;120(20):4123–33.

44. Douglas NM, Pontororing GJ, Lampah DA, Yeo TW, Kenangalem E, Poespoprodjo JR, et al. Mortality attributable to Plasmodium vivaxmalaria: a clinical audit from Papua, Indonesia. BMC Medicine. 2014;12(1):217. doi: 10.1186/s12916-014-0217-z.

45. Abraham P, Lubis IND, Novivanti R, Trianty L, Nisa FA, Kariodimedjo P, et al. The household economic burden of human-only and zoonotic malaria, compared to other causes of acute febrile illness in Indonesia. BMJ Global Health. 2026;11(3).

46. The World Bank. Implicit GDP deflator (base year = 2010 = 100) [NY.GDP.DEFL.ZS.AD] 2025. Available from: https://data.worldbank.org/indicator/NY.GDP.DEFL.ZS.AD.

47. Bank TW. Official exchange rate (LCU per US$, period average) [PA.NUS.FCRF] 2025. Available from: https://data.worldbank.org/indicator/PA.NUS.FCRF.

48. Statistics Indonesia. Kajian Lifetable Indonesia. Berdasarkan Hasil SP2010. Studies Lifetable Indonesia Based on results of the 2010 Population Census. 2015.

49. Indonesian Health Technology Assessment Committee. General Guideline for Health Technology assessment in Indonesia. In: The Centre for Health Financing and Decentralization Policy MoH, editor. 2022.

50. The World Bank. GDP per capita (current US$ - Indonesia) 2025. Available from: https://data.worldbank.org/indicator/NY.GDP.PCAP.CD?locations=ID.

51. Briggs A, Fenn P. Confidence intervals or surfaces? Uncertainty on the cost-effectiveness plane. Health economics. 1998;7(8):723–40.

52. Fransisca L, Ome-Kaius M, Laman M, Poespoprodjo JR, Pasaribu AP, Sutanto I, et al. High-dose, short-course primaquine after point-of-care G6PD testing for the radical cure of Plasmodium vivax malaria: a safety study in Papua New Guinea and Indonesia. The Lancet Regional Health–Western Pacific. 2026;71.

53. Drake TL, Lubell Y. Malaria and economic evaluation methods: challenges and opportunities. Applied health economics and health policy. 2017;15(3):291–7.

54. Shretta R, Avanceña AL, Hatefi A. The economics of malaria control and elimination: a systematic review. Malaria Journal. 2016;15(1):593.

55. Shretta R, Silal SP, Celhay OJ, Mercado CEG, Kyaw SS, Avancena A, et al. Malaria elimination transmission and costing in the Asia-Pacific: Developing an investment case. Wellcome Open Research. 2020;4:60.

56. Sarma N, Patouillard E, Cibulskis RE, Arcand J-L. The economic burden of malaria: revisiting the evidence. The American journal of tropical medicine and hygiene. 2019;101(6):1405.

57. Mezieobi KC, Alum EU, Ugwu OP-C, Uti DE, Alum BN, Egba SI, et al. Economic burden of malaria on developing countries: a mini review. Parasite Epidemiology and Control. 2025;30:e00435.

58. Hanandita W, Tampubolon G. Geography and social distribution of malaria in Indonesian Papua: a cross-sectional study. International journal of health geographics. 2016;15(1):13.

59. Ciavarella C, Drakeley C, Price RN, Mueller I, White M. Quantifying Plasmodium vivax radical cure efficacy: a modelling study integrating clinical trial data and transmission dynamics. The Lancet Infectious Diseases. 2025;25(6):668–77.

60. Indonesia PSB. Standard M 10 System (Module) — E–Catalogue (LKPP, Indonesia) 2025. Available from: https://e-katalog.lkpp.go.id/katalog.produkctr/getdetailproductcenter?id=2772845.

61. MedAccess. G6PD testing 2024. Available from: https://medaccess.org/our-agreements/agreements/g6pd-testing/.

